# Functional and Structural Neuroanatomic Correlates of Intraoperative Anesthetic Dose-Adjusted EEG Alpha Power Among Older Surgical Patients

**DOI:** 10.64898/2026.10.21.26366127

**Authors:** Tyler Reekes, Jeffrey N. Browndyke, Melody Reese Smith, Yotam Weiss, Jian Yu, Mary Cooter Wright, Hayden Kang, Noah J Timko, Piper Boykin, Bethany Hsia, Michael J. Devinney, Miles Berger

**Affiliations:** Department of Anesthesiology, Duke University Medical Center, Durham, NC, USA; Center for the Study of Aging and Human Development, Duke University Medical Center, Durham, NC, USA; Department of Anesthesiology, Perioperative and Pain Medicine, Stanford School of Medicine, Stanford, CA, USA; Duke-University of North Carolina (UNC) Alzheimer’s Disease Research Center, Durham, NC, USA; Department of Psychiatry and Behavioral Sciences, Duke University Medical Center, Durham, NC, USA; Duke Institute for Brain Sciences, Duke University, Durham, NC, USA

**Author notes:** These authors contributed equally to this manuscript and are co-first authors (Tyler Reekes, Jeffrey N. Browndyke). These authors contributed equally to this manuscript and are co-senior authors (Michael J. Devinney, Miles Berger). Corresponding Author: Miles Berger, MD, PhD, Department of Anesthesiology, Perioperative and Pain Medicine, Stanford University, 300 Pasteur Dr, Room H3580, MC 5640, Stanford CA 94305.

## Abstract

**Background:** Alterations in intraoperative electroencephalography (EEG) alpha power and anesthetic dose-adjusted EEG alpha power (ADA-FP alpha power) have been associated with both preoperative cognitive impairment and postoperative delirium (POD) among older surgical patients, yet the neurologic underpinnings of alterations in these anesthetic brain response patterns are unknown.

**Methods:** We conducted a single center prospective observational cohort study (ALADDIN), of older adults undergoing non-cardiac/non-cranial surgery, which included preoperative structural and resting state functional MRI, lumbar punctures to obtain CSF, 32 channel intraoperative EEG recordings, and repeated POD assessments. To account for anesthetic dose, frontoparietal EEG alpha power (ADA-FP alpha power) was adjusted using our established formula: [intraoperative alpha power]/(2.5 - end-tidal MAC fraction). Multivoxel pattern analysis (MVPA) with follow-up seed-to-voxel analyses identified functional connectivity patterns associated with intraoperative ADA-FP alpha power. Voxel-wise regression analysis with threshold free cluster estimation (TFCE) correction was used to evaluate associations between cortical thickness and ADA-FP alpha power. CSF biomarker values were obtained from NULISA proximity extension assays. Delirium incidence and severity was assessed with the 3-Minute Diagnostic Confusion Assessment Method (3D-CAM). Delirium severity area under the curve (AUC) during hospitalization was defined as the sum of all 3D-CAM delirium severity scores within the postoperative hospitalization period (for up to 7 days after surgery). The Confusion Assessment Method for the Intensive Care Unit (CAM-ICU) was used in cases of non-verbal/intubated patients.

**Results:** ALADDIN enrolled 215 patients, of whom 168 underwent surgery and had high-quality intraoperative EEG data (44.6% female; mean age 70.9 years). MVPA showed that ADA-FP alpha power was associated with altered functional connectivity within the bilateral frontal poles, left superior frontal gyrus and the right superior parietal lobule (p-FWE < 0.05). Seed-to-voxel analyses revealed that higher ADA-FP alpha power was associated with increased connectivity between DMN MVPA regions and DMN, DAN and salience/ventral attention and decreased connectivity with visual regions. Conversely, right superior parietal lobule connectivity shifted towards greater DMN segregation and increased somatosensory/motor coupling (p-FWE < 0.05). Lower ADA-FP alpha power was also associated with lower cortical thickness in left occipitotemporal and superior parietal cortices (TFCE-corrected p < 0.05). ADA-FP alpha power was also lower among participants with (vs those without) postoperative ward/floor delirium (mean difference, 1.33; 95% CI, 0.22 to 2.44; p=0.019), and ADA-FP alpha power was inversely associated with peak delirium severity (spearman’s ρ=−0.26; 95% CI, −0.40 to −0.11; p <0.001) and delirium severity AUC (spearman’s ρ=−0.28; 95% CI, −0.42 to −0.13; p=<0.001).

**Discussion:** These findings suggest a functional and structural neuroanatomic basis for the decreased EEG ADA-FP alpha power values seen among some older surgical patients, which may explain the association between decreased EEG ADA-FP alpha power and increased rates of both preoperative cognitive impairment and postoperative delirium. More specifically, these findings suggest that altered default mode and dorsal attention network connectivity is linked to decreased EEG ADA-FP alpha power and raise the possibility that preoperative/baseline functional connectivity alterations within these networks may play a role in delirium risk.

## Introduction

Postoperative delirium (POD) is an acute disturbance in attention, awareness, and cognition. Although its incidence varies considerably across surgical populations, settings, and hospitals, rates as high as 50% continue to be reported.(1) POD is associated with both short- and long-term adverse outcomes, including mortality, institutionalization, and neurocognitive disorders such as dementia, and imposes a substantial economic burden.(2) These consequences have established POD as an important target for perioperative prevention.(3) However, its multifactorial etiology, reflecting the interaction between preexisting vulnerabilities and perioperative stressors, makes it difficult to precisely identify vulnerable patients before POD becomes clinically apparent.(4) Because POD is a fluctuating syndrome, binary incidence alone may incompletely capture its clinical expression; timing, peak severity, and cumulative burden may provide complementary characterization of its clinical course and burden.(5–7)

Postoperative delirium risk is also known to be increased among patients who have decreased intraoperative electroencephalography (EEG) power within the 8-12 Hz range(8–10), known as alpha power. Normally, inhaled anesthetics induce prominent frontal and parietal EEG alpha oscillations, (11) while lower anesthesia-induced EEG alpha power has been associated with worse preoperative cognitive function, suggesting that the brain’s electrophysiologic response to anesthesia may provide a physiologic probe of neurocognitive function.(10) Subsequent studies have shown that, independent of age, lower alpha power is also associated with an increased propensity for EEG burst suppression, an EEG pattern characterized by intermittent bursts of activity alternating with periods of suppressed near-isoelectric activity.(12) Like alpha power, burst suppression has also been associated with preoperative cognitive impairment and increased postoperative delirium risk, as well as with postoperative mortality, and discharge to long-term care. (8–10,13,14). Taken together, these findings support low intraoperative alpha power as a marker of a vulnerable brain and of increased risk for multiple adverse postoperative outcomes.

These converging associations between low intraoperative alpha power (increased burst suppression) with lower baseline cognition and worse postoperative outcomes support the concept of anesthesia as a neurophysiologic stress test of the aging brain.(15) Rather than serving as a traditional measure of anesthetic depth, the EEG response to anesthetic exposure may thus reveal latent differences in brain functional capacity or reserve that are not apparent from baseline measures alone.

Prior work from our group characterized the relationship between age-adjusted end-tidal minimum alveolar concentration (aaMAC) and the EEG response to volatile anesthetics, and used this framework to develop a novel measure of brain anesthetic resistance.(16) This approach uses 2.5 aaMAC as a reference ceiling, representing the highest aaMAC likely to be observed in routine adult clinical anesthesia practice.(16) Building on this framework, anesthetic dose-adjusted frontoparietal alpha power (ADA-FP alpha power) was developed to account for differences in anesthetic exposure. In a pooled analysis of three prospective studies, lower ADA-FP alpha power was associated with both POD and worse preoperative performance on timed tests of processing speed and executive function.(17)

To investigate the brain basis of this EEG phenotype, magnetic resonance imaging (MRI) can provide both structural characteristics such as regional brain morphology, cortical thickness and tissue integrity, and functional information to identify patterns of coordinated activity between brain regions that are not necessarily anatomically connected. Resting state fMRI can characterize intrinsic connectivity of within spatially distant brain networks, including the default mode and dorsal attention networks..(18–21)

Prior perioperative work has linked changes in resting-state connectivity within default mode network (DMN) regions to postoperative cognitive change in older adults.(22) However, the structural and functional brain basis of reduced intraoperative alpha power remains unknown. To our knowledge, no prior study has examined whether preoperative structural and functional neuroimaging phenotypes are associated with intraoperative ADA-FP alpha power (or intraoperative alpha power or burst suppression without adjustment for anesthetic dose). Thus, we designed the Low neurophysiologic resistance to <u>A</u>nesthetics as a marker of preclinical/prodroma<u>L A</u>lzheimer’s <u>D</u>isease and neurovascular pathology, <u>D</u>elirium risk and <u>IN</u>attention (ALADDIN) study to address this question by relating preoperative cortical thickness and functional connectivity measures to subsequent intraoperative ADA-FP alpha power within the same individuals. In effect, this allows us to examine whether the electrophysiologic response to the anesthesia and surgery “stress test”(15) reflects underlying structural brain features, changes in functional brain network connectivity, or both.

## Methods

### Study design and participants

The ALADDIN study was approved by the Duke University Health System institutional review board prior to study initiation. Written informed consent was obtained from all participants before study participation, and all study procedures were conducted in accordance with the Declaration of Helsinki.

Participants were identified through operating room schedule screening and contacted by study personnel by telephone or electronic health record messaging. Inclusion criteria included adults aged ≥60 years undergoing non-cardiac and non-intracranial surgery scheduled for at least 120 minutes with a planned postoperative hospitalization for at least 1 day. Exclusion criteria included signs or symptoms of systemic infection, receipt of chemotherapeutic agents known to impair cognition, inability to safely undergo the study lumbar puncture, and a history of malignant hyperthermia. Participants also underwent standard MRI safety screening, and those unable to undergo MRI because of safety contraindications or claustrophobia were excluded. Study procedures included preoperative neurocognitive assessments, lumbar punctures to obtain cerebrospinal fluid (CSF) samples, preoperative functional and structural MRI scans, intraoperative EEG recording and POD assessments.

### Neurocognitive assessment

Preoperative global cognitive performance was assessed using the Montreal Cognitive Assessment (MoCA) screening tool.

### Lumbar Punctures, CSF processing, and NULISA Biomarker Assay

CSF samples were collected via lumbar puncture using a 25g pencil-point needle, following our established protocol to reduce pain and complications, and aliquoted and stored at -80°C in Sarstedt microcentrifuge polypropylene tubes (VWR; Radnor, PA).(23) Biomarker profiling was performed by the Human Immune Monitoring Center (HIMC) at Stanford University using the NULISAseq Inflammation Panel 250 (Alamar BioSciences, Fremont, CA, USA), a multiplexed proximity ligation assay targeting 250 inflammation-associated proteins. Samples were processed automatically using the ARGO HT system (Alamar BioSciences). Twenty-five microliters of each CSF sample were loaded onto the sample plate together with three sample controls, four negative controls, and three inter-plate controls. Following completion of the automated assay, next-generation sequencing of the pooled library was performed using the AVITI platform (Element Biosciences, San Diego, CA, USA). Data were generated using Alamar Command Center and NULISA Analysis Software, with normalization to internal controls and inter-plate controls. Values were subsequently log2-transformed and reported in NULISA Protein Quantification (NPQ) units.

### Preoperative structural and functional MRI acquisition and preprocessing

Preoperative MRI was performed on a 3T GE Signa MR750 MRI scanner (GE Healthcare, Waukesha, WI, USA) equipped with a 48 channel receive-only head coil, at the Duke Brain Imaging and Analysis Center. Anatomical data consisted of a high-resolution T1-weighted axial oblique 3D Magnetization-Prepared Rapid Gradient-Echo (MP-RAGE) with TR 2.24 s, TE 2.9 ms, TI 900 ms, flip angle 8°, 256 x 256 matrix, and 1mm slice thickness. Rs-fMRI data were acquired using the Inscapes paradigm(24) using a gradient-echo, echo-planar imaging (EPI) sequence with a TR of 1500 ms, TE 30 ms, flip angle 80°, 128 x 128 matrix, 2 mm slice thickness, 66 slices, and a multiband acceleration factor of 3. For the majority of participants (97%) preoperative assessments were performed < 6 weeks surgery (median = 8 days; range 1-141 days). The remaining 3% had extended delays between timepoints due to rescheduled surgeries or other scheduling conflicts. T1-weighted images were processed using Statistical Parametric Mapping (SPM12) and the Computational Anatomy Toolbox (CAT12) for tissue segmentation and surface- and volume-based morphometric analyses, including regional cortical thickness and cortical volume.(25)

Preprocessing and analysis of fMRI data were performed using CONN(26) release 22.v2407(27) and SPM(28) release 25.01.02. Functional and anatomical data were preprocessed using a standardized, modular preprocessing pipeline(29) including removal of initial scans, realignment with correction of susceptibility distortion interactions, slice timing correction, outlier detection, direct segmentation and MNI-space normalization, and smoothing. The first 12 scans (18 seconds) in each functional run were removed to avoid magnetic field inhomogeneities. Functional data were realigned using the SPM realign & unwarp procedure(30), where all scans were co-registered to a reference image (first scan of the session) using a least squares approach and a 6 parameter (rigid body) transformation(31), and resampled using b-spline interpolation to correct for motion and magnetic susceptibility interactions. Temporal misalignment between different slices of the functional data was corrected using the SPM slice-timing correction (STC) procedure(32,33), with sinc temporal interpolation to resample each slice Blood-Oxygen-Level-Dependent (BOLD) timeseries to a common mid-acquisition time. Potential outlier scans were identified using Artifact Recognition Tool (ART)(34) as acquisitions with framewise displacement values above 0.5 mm or global BOLD signal changes more than 5 standard deviations from the mean (35,36). A reference BOLD image was computed for each subject by averaging all scans excluding outliers. Functional and anatomical data were normalized into standard MNI space, segmented into grey matter, white matter, and CSF tissue classes, and resampled to 2 mm isotropic voxels following a direct normalization procedure(36,37) using a SPM unified segmentation and normalization algorithm(38,39) with the default IXI-549 tissue probability map template. Last, functional data were smoothed using spatial convolution with a Gaussian kernel of 8 mm at full width half maximum (FWHM).

In addition, functional data were denoised using a confound regression pipeline incorporating established nuisance regressors.(29) This included the removal of potential confounding effects derived from white matter (5 regressors), CSF (5 regressors), motion parameters and their first order derivatives (12 regressors)(40), session effects and their first order derivatives (2 regressors), and linear trends (2 regressors) within each functional run, followed by bandpass frequency filtering of the BOLD timeseries(41) between 0.008 Hz and 0.09 Hz. CompCor regressors(42,43) were estimated within each subject’s eroded white matter and CSF masks by extracting the mean signal and the largest principal components orthogonal to the mean signal, motion parameters, and outlier scans. Participants were excluded from functional MRI analyses if they had less than 9 minutes of valid resting state functional connectivity data per established recommendations.(44)

### MRI data analysis

#### Multivoxel Pattern Analysis (MVPA) of relationships between baseline/preoperative Resting state functional connectivity and intraoperative ADA-FP alpha power

To identify brain regions whose functional connectivity varied in association with ADA-FP alpha power, we used multivoxel pattern analyses (MVPA)(45). Whole-brain functional connectivity across subjects’ were computed from the matrices of bivariate correlation coefficients between the BOLD timeseries from each pair of voxels, estimated via a singular value decomposition of the z-score normalized BOLD signal (subject-level SVD) with 64 components separately for each subject.(29)

Eigenpatterns and eigenpattern-scores were then computed separately for each individual seed voxel as the left- and right-singular vectors, respectively, from a singular value decomposition (group-level SVD) of the matrix of functional connectivity values between this seed voxel and the rest of the brain (a matrix with one row per target voxel, and one column per subject). From these eigenpatterns, 6 associated eigenpattern-score images were derived for each individual subject characterizing their brain-wide functional connectome state.

The six eigenpattern scores were entered as dependent variables in a whole-brain, voxel-wise MVPA regression model with ADA-FP alpha power as the primary regressor with age and mean in-scanner motion entered as covariates. Voxel-level hypotheses were evaluated using multivariate parametric statistics with random-effects across subjects and sample covariance estimation across multiple measurements. Inferences were performed at the level of individual clusters (groups of contiguous voxels). Cluster-level inferences were based on parametric statistics from Gaussian Random Field theory.(29,46) Results were thresholded using a combination of a cluster-forming p < 0.001 voxel-level threshold, and a familywise corrected p-FWE < 0.05 cluster-size threshold.(47)

### Seed-to-Voxel Analysis

Follow-up seed-to-voxel regression analyses were conducted for each significant cluster identified by the primary MVPA analysis using the same model parameters and statistical thresholds to identify contributory foci, and to assess the directionality of associations between dose-adjusted FP alpha power and resting state functional connectivity between each given seed and the contributory foci.

### Cortical thickness analysis

Cortical thickness was estimated using CAT12. Voxel-wise regression analysis was performed in SPM12 to examine the association between dose-adjusted FP alpha power with age included as a covariate. Significance was assessed using threshold free cluster enhancement (TFCE) to account for multiple comparison correction across voxels. Significance was set at p-TFCE < 0.05.

### Intraoperative EEG acquisition and preprocessing

Intraoperative EEG data were recorded using a tethered 32-electrode gel-based cap (BrainAmp MR Plus, Brain Products GmbH, Gilching, Germany) and wireless Bluetooth LiveAmp amplifier (LiveAmp, Brain Products GmbH, Gilching, Germany). Prior to data collection, electrode impedances generally < 50 kΩ were obtained by light abrasion of the scalp at each electrode location with coarse conductive paste (e.g., Abralyt HiCl 1000, EASYCAP GmbH, Herrsching, Germany). EEG signals were recorded at a native sampling rate ≥ 500 Hz with a 0.016-250 Hz band-pass filter.

Offline EEG processing was performed in MATLAB R2023a (The MathWorks, Inc., Natick, MA, USA) using EEGLAB v2025.0.0 (Swartz Center for Computational Neuroscience, San Diego, CA, USA) and custom scripts. Raw EEG data were bandpass filtered between 0.5 and 55 Hz using a Hamming-windowed FIR filter (order 3300) and downsampled to 250 Hz. Artifact detection was performed on 1-second non-overlapping windows using three criteria. (1) Windows were flagged as artifact if the signal variance fell below 0.0001 µV² (an SD < ∼0.01 µV, indicative of poor electrode contact) or above 1,000 µV² (an SD > ∼32 µV, indicative of large-amplitude artifacts). (2) Windows were also flagged as artifact if the absolute value of the mean amplitude exceeded 100 µV (indicative of non-neural signals). (3) Channel-specific dynamic outlier detection thresholds were also applied to remove extreme values based on the bounds of each patient’s typical intraoperative EEG signal. For each electrode, 1-second windows exceeding 3 robust standard deviations (SDs), which are approximately 1.4826 times the median absolute deviation, above the median variance across the recording were flagged as artifact. This is similar to traditional outlier detection for values 3 SDs beyond the mean, although, unlike mean and SD, median and robust SD are more resistant to bias from extreme values such as amplitude deflections induced by surgical movement or electrocautery. Thus, the 3 robust SD approach defines a unique, patient-specific threshold for detection of outliers that might otherwise be missed by absolute thresholds, given known between-patient variability in underlying brain activity and intraoperative recording conditions (e.g., anesthetic depth, surgical invasiveness, patient repositioning). For any epoch flagged as artifact by any of the above criteria, we also excluded a conservative 5-second buffer of EEG data on each side to account for potential artifact seepage into adjacent EEG data.

These analyses focused on frontal and parietal electrode subsets. Thus, regions of interest (ROIs) included the average signal from electrode sites Fp1 + F3 + F7 (a left frontal ROI), Fp2 + F4 + F8 (a right frontal ROI), Fz (a central frontal ROI), P3 + P7 (a left parietal ROI), P4 + P8 (a right parietal ROI), and Pz (a central parietal ROI), which combine to represent a larger, combined frontoparietal region. Seconds of preprocessed EEG data were marked as unusable and replaced with missing values when all channels within a given ROI were simultaneously flagged as artifactual by any of the three preprocessing criteria. When only some ROI channels were flagged as artifactual, the ROI average was computed with the flagged channels contributing zero power. Once preprocessed, the EEG data were re-referenced to the average signal of the TP9 and TP10 electrodes near the mastoid bone (i.e., standard 10–20 left and right mastoid electrode sites), as previously reported(10,17), or to either TP9 or TP10 when only one mastoid electrode signal passed preprocessing criteria.

### EEG post-processing and intraoperative FP alpha power calculation

After preprocessing and re-referencing, intraoperative FP alpha power was computed with Chronux multitaper (mtspecgramc) using a 10 second window, 1 second step, and 5 discrete tapers with a time-bandwidth product of 3, corresponding to ∼0.6 Hz full spectral smoothing. Linear power was averaged across channels within each ROI per second, then across seconds to minutes (when there was at least one second of non-missing data), across ROIs for the combined FP region (when there was non-missing data from at least one frontal and one parietal ROI), integrated over 7-13Hz, and then converted to decibels (dB). To avoid EEG signal fluctuations during induction and emergence from the anesthetized state, we analyzed ADA-FP alpha power starting from > 5 min after anaesthesia began until > 5 min before the case ended (based on electronic medical record anaesthesia start and stop time notes). Minutes of artifact were marked as missing data and minutes without valid age adjusted MAC (aaMAC) data (such as during non-volatile anaesthetic use) were excluded from this analysis. To minimise potential confounding, EEG data were also excluded during case minutes corresponding to extreme aaMAC doses (< 0.5 or ≥ 2.5), during nitrous oxide or lidocaine administration, during and for 30 min after ketamine or dexmedetomidine administration, and during and for 10 minutes post-propofol infusion or bolus, because of the differential effects of these drugs on the EEG vs those of volatile anaesthetics. Patients with fewer than 10 minutes that survived all minute-level exclusions (timing buffers, aaMAC criteria, and drug windows) were excluded from further analyses. Because FP alpha power required data availability from at least one frontal and at least one parietal ROI, FP alpha power could be missing during a minute with otherwise analyzable EEG data. Thus, some patients could have < 10 minutes of available FP alpha power data.

### Anesthetic dose-adjustment of intraoperative frontoparietal alpha power

To account for interindividual differences in anesthetic exposure, FP alpha power was adjusted for the concurrent volatile anesthetic dose on a minute-by-minute basis using the previously described formula: (17)

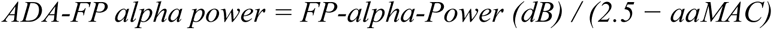

where aaMAC represents the age-adjusted end-tidal minimum alveolar concentration fraction of volatile agents at the corresponding time point, and 2.5 represents the maximum theoretical aaMAC value that a patient typically receives during anesthesia. Minute-level dose-adjusted values were summarized as the mean of median values over a sliding 5-min window (with 4-min overlap, minimum of 1 valid minute per window) to minimize the impact of any remaining artifact across the surgical case. This mean of median filtering generates a single summary ADA-FP alpha power value for each patient across their entire anesthetic case.

### Postoperative delirium screening

Delirium was assessed preoperatively and postoperatively in the post-anesthesia care unit (PACU), on the evening of the day of surgery, and twice daily (morning and evening) during hospitalization for up to 7 days or until discharge, using the 3-Minute Diagnostic Confusion Assessment Method (3D-CAM) or the Confusion Assessment Method for the Intensive Care Unit (CAM-ICU) for intubated patients. Delirium assessments were performed by study personnel trained by a licensed clinical neuropsychologist (JNB) using materials from the Hebrew SeniorLife/Harvard Medical School Successful Aging after Elective Surgery (SAGES) study group.(48) Participants with at least one positive 3D-CAM or CAM-ICU assessment were considered positive for POD. For sensitivity analyses of delirium timing, participants were classified as having no POD at any time point, delirium only detected during the patient’s post-anesthesia care unit stay (PACU delirium), or postoperative delirium that occurred on the hospital ward/floor after the patient’s discharge from the PACU (ward POD), regardless of whether the patient experienced PACU POD.(7)

Delirium severity was measured on a 0-20 point scale from the 3D-CAM as previously described.(5) Cumulative delirium burden was quantified as the area under the curve (AUC) of delirium severity over time during the patients’ postoperative hospitalization (up to 7 days following surgery).(6)

### Comparison between ADA-FP alpha power and postoperative delirium metrics

ADA-FP alpha power was first compared between participants with and without POD using an independent-samples t-test. In a sensitivity analysis examining delirium timing, separate independent-samples t-tests compared ADA-FP alpha power between participants with no POD and those with PACU-only POD, and between participants with no POD and those with hospital ward POD. Because delirium severity and cumulative delirium burden were non-normally distributed, associations between ADA-FP alpha power and peak delirium severity and delirium severity-time AUC were evaluated using Spearman rank correlations.

Continuous variables were summarized using means ±SD or medians and interquartile ranges (IQR), as appropriate, and categorical variables as counts and percentages (n%). All tests were two-sided, with P<0.05 considered statistically significant.

## Results

### Cohort characteristics

A total of 215 participants were enrolled in ALADDIN, 168 of whom had adequate intraoperative EEG ADA-FP alpha power for analysis (Figure 1). Demographic/baseline and perioperative patient characteristics are described in Table 1 and Supplementary Table 1, respectively. Overall, 44.6% of participants were female and 12.5% identified as Black or African American. Mean age was 70.9 years (± 5.0), with median education of 15 years [12.8, 18] and the median baseline MoCA score was 25 [21, 26]. No participant was positive for delirium at baseline by 3D-CAM. Mean surgical duration was 214 minutes (± 132) or ∼3.5 hours. Mean aaMAC was 0.9 (± 0.2) and estimated blood loss was 97 mL (± 313; Supplemental Table 1).

**Figure 1.**
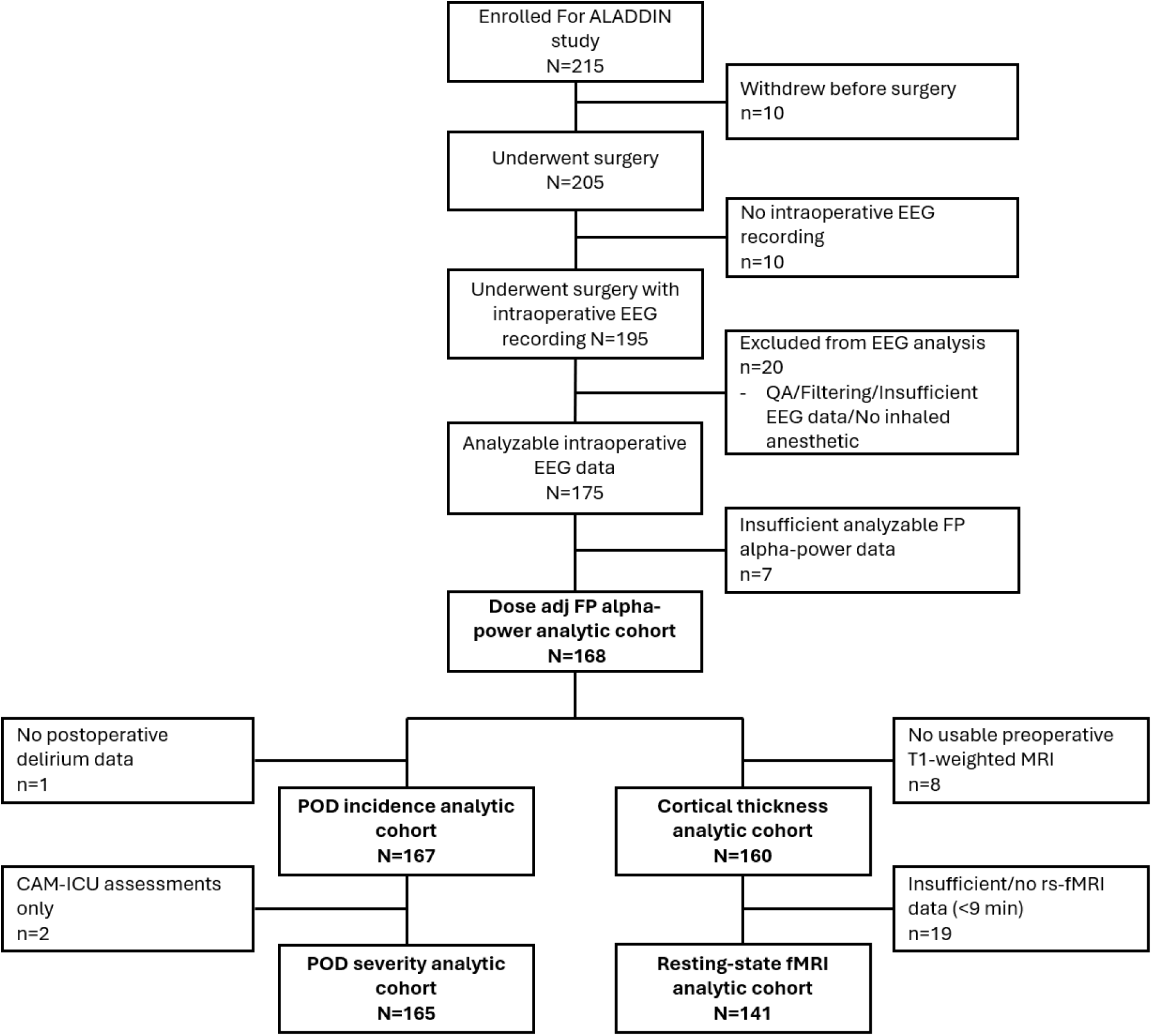
STROBE Diagram of Participant Enrollment. Participant flow from enrollment through modality-specific analytic cohorts. The dose-adjusted frontoparietal (FP) alpha-power analytic cohort comprised participants with sufficient analyzable intraoperative EEG data and volatile anesthetic exposure. Delirium incidence and severity analyses were restricted according to availability of POD assessments and severity scores, respectively. MRI analytic cohorts were defined independently from the dose-adjusted FP alpha-power cohort according to availability and quality of preoperative T1-weighted MRI and resting-state functional MRI (rs-fMRI) data. CAM-ICU, Confusion Assessment Method for the Intensive Care Unit; EEG, electroencephalography; FP, frontoparietal; MRI, magnetic resonance imaging; POD, postoperative delirium; rs-fMRI, resting-state functional magnetic resonance imaging.

**Table 1:** Baseline Cohort/Patient Characteristics.

|  |  |
| --- | --- |
|  | n = 168 |
| Age | 70.9 (5.0) |
| Sex |  |
| Male | 93 (55.4%) |
| Female | 75 (44.6%) |
| Race |  |
| Caucasian/White | 140 (83.3%) |
| Black/African American | 21 (12.5%) |
| Asian | 2 (1.2%) |
| Alaskan/Pacific Islander | 2 (1.2%) |
| Multiracial | 2 (1.2%) |
| Unknown | 1 (0.6%) |
| BMI | 27.3 [25, 30.7] |
| Height (cm) | 173 [163, 180] |
| Weight (kg) | 82.3 [69.3, 93.9] |
| MoCA | 25 [21, 26] |
| VWR | 4 [0, 7.25] |
| Delirium at baseline | 0 |

Data are presented as n (%), mean (SD), or median [25th, 75th percentiles], as appropriate. BMI, body mass index; MoCA, Montreal Cognitive Assessment; VWR, verbal word recall.

Among participants with usable EEG, 141 had sufficient preoperative rs-fMRI data without excessive motion (>9 min) and were included in the primary MVPA analysis. For cortical thickness analysis, 160 of the 168 participants with adequate EEG data also had usable T1 anatomical data.

### Preoperative functional connectivity patterns are associated with ADA-FP alpha power

Whole-brain MVPA controlling for age and mean motion identified four spatially distinct clusters in which preoperative resting-state functional connectivity patterns were significantly associated with intraoperative dose-adjusted FP alpha power (FWE-corrected p<0.05; Figure 2). These clusters were located within the left and right frontal poles, the left superior frontal gyrus, and the right superior parietal lobule (Table 2). We then used the Schaefer et al. 7-network (400 ROIs) atlas (49) to characterize which functional networks each of these four MVPA clusters belonged to, which demonstrated that the left and right frontal poles and the left superior frontal gyrus belong to the default mode network, while the right superior parietal lobule cluster belongs to the cognitive control network/dorsal attention network (Table 2).

**Figure 2.**
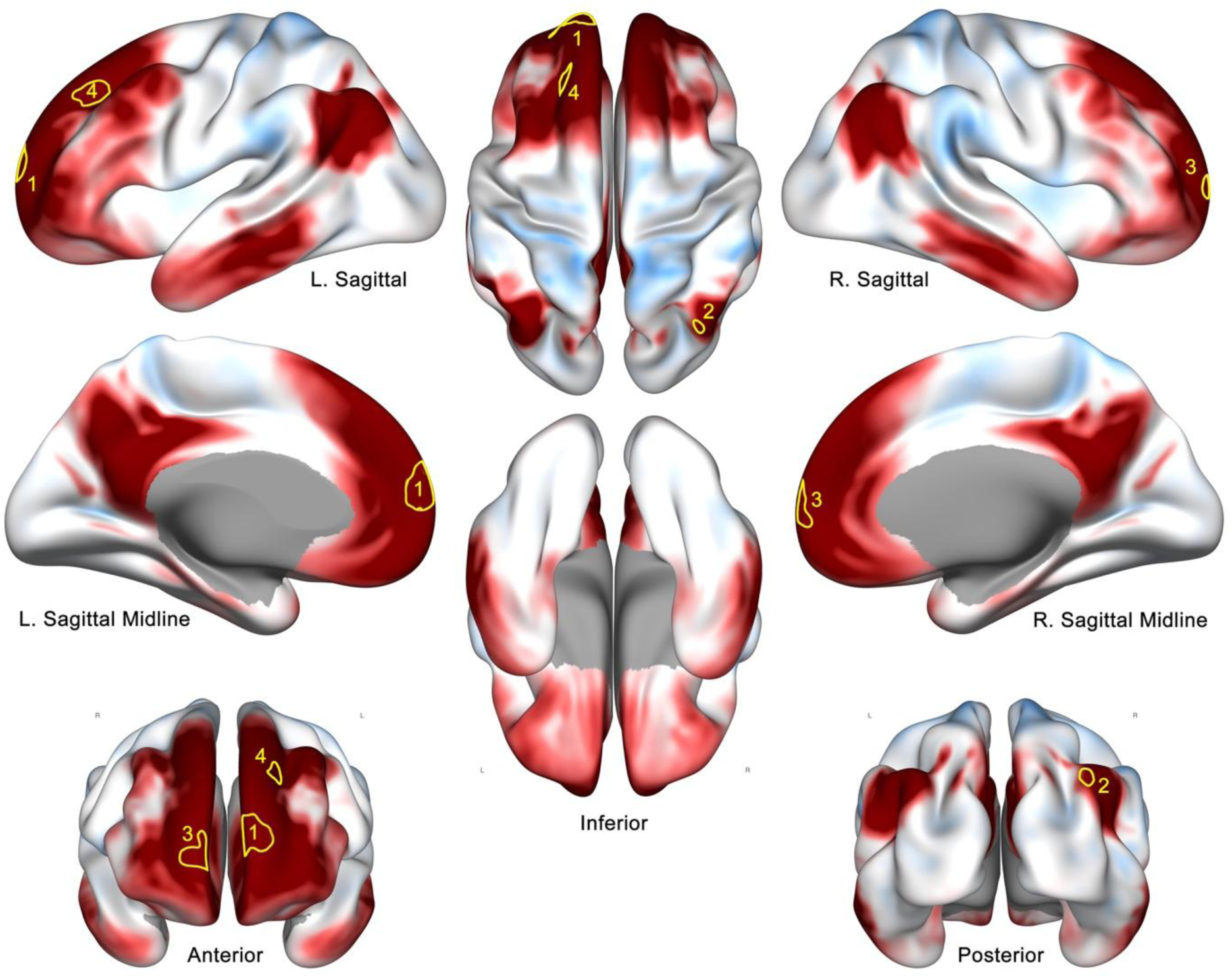
Multivoxel Pattern Analysis (MVPA) of associations between Preoperative Resting State Functional Connectivity and Intraoperative Anesthetic-Dose Adjusted FrontoParietal EEG Alpha Band Power

**Table 2:** Multi-Voxel Pattern Analysis (MVPA) cluster statistics.

| MVPA cluster | Anatomical Location | Cluster maxima (x, y, z) | Cluster extent (k) | Cluster p-FWE | Functional Association |
| --- | --- | --- | --- | --- | --- |
| 1 | Left Frontal Pole | (-8, 66, 10) | 244 | < 0.001 | Default Mode Network |
| 2 | Right Superior Parietal Lobule | (22, -72, 50) | 69 | 0.018 | Cognitive Control/<br>Dorsal Attention |
| 3 | Right Frontal Pole | (6, 64, 0) | 68 | 0.019 | Default Mode Network |
| 4 | Left Superior Frontal Gyrus | (-14, 38, 38) | 59 | 0.04 | Default Mode Network |

Next, to evaluate the possibility that the relationships between preoperative resting state functional connectivity within these brain regions (described above) and with intraoperative dose-adjusted FP alpha power were driven by preoperative amyloid pathology, we repeated the MVPA analyses described above while controlling for the CSF Aβ42/Aβ40 ratio (Supplementary Figure 1) and again while controlling for CSF ptau-217 levels (Supplementary Figure 2). In both these sensitivity analyses, similar brain regions were identified as showing correlations between preoperative resting state functional connectivity and intraoperative dose-adjusted FP alpha power as in our original analysis (Figure 2). This indicates that the correlations between resting state functional connectivity within the left and right frontal poles, left superior frontal gyrus, and right superior parietal lobule and intraoperative dose-adjusted FP alpha power are not driven by or confounded by baseline/preoperative amyloid pathology.

Next, we performed follow-up seed-to-voxel analysis to identify brain regions whose functional connectivity strength to these MVPA seeds varied in association with ADA-FP alpha power, and to identify the directionality of these associations. Higher alpha power was generally associated with greater connectivity between MVPA seeds within the DMN and other DMN, DAN and salience/ventral attention (VAN) regions and inversely associated with connectivity to visual network regions (Figure 3 panels B.1, 3 and 4). In contrast, higher alpha power was associated with a shift toward weaker or more negative connectivity between the MVPA region within the right superior parietal lobule (a hub of the cognitive control network/DAN) and regions of the default mode network, alongside stronger or more positive connectivity with region of the somatosensory/motor network (Figure 3 panel B.2, Supplementary Table 2). This patten may reflect greater functional segregation between networks responsible for internally versus externally directed attention.

**Figure 3.**
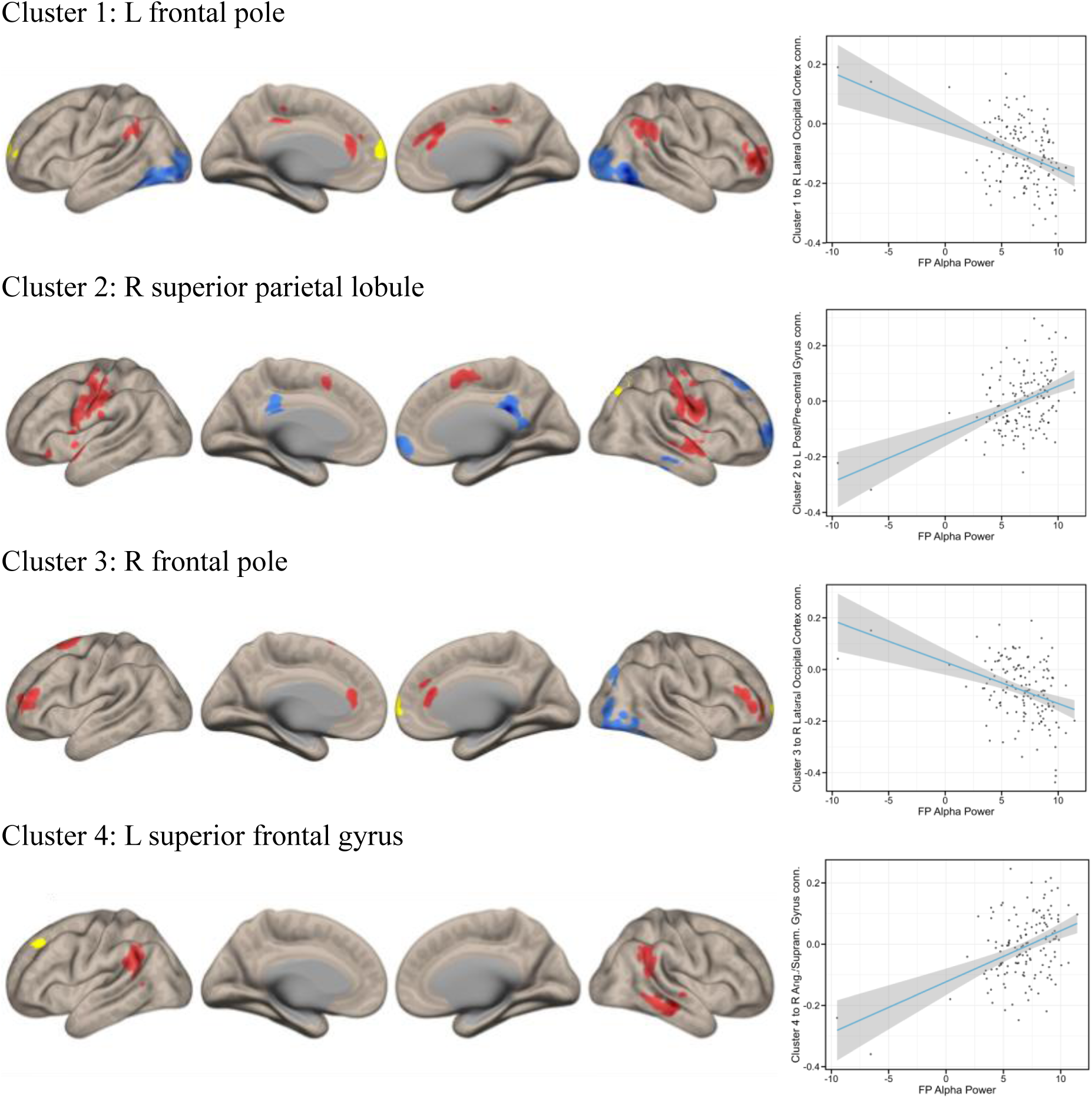
Seed-to-voxel functional connectivity associations with intraoperative anesthetic dose-adjusted frontoparietal alpha power. Region color indicates the direction of the association between Anesthetic-Dose Adjusted FrontoParietal Alpha Power and functional connectivity between the seed region (MVPA cluster, indicated in yellow) and the resultant seed-to-voxel clusters. Red indicates a positive relationship, and blue indicates a negative relationship between functional connectivity strength and Anesthetic-Dose Adjusted FrontoParietal Alpha Power.

### Lower ADA-FP alpha power is associated with reduced cortical thickness

Regional cortical thickness analyses showed a predominantly left-lateralized pattern of association with ADA-FP-alpha-Power. Lower ADA-FP-alpha-Power was associated with cortical thinning in the left temporoparietal and temporal cortices, including the angular/inferior parietal region, posterior superior and middle temporal cortex, and ventral temporal regions encompassing the fusiform/parahippocampal cortex. White outlines in Figure 4 reflect regions in which lower ADA-FP-alpha-Power was associated with cortical thinning at a TFCE-corrected p<0.05 threshold.

**Figure 4.**
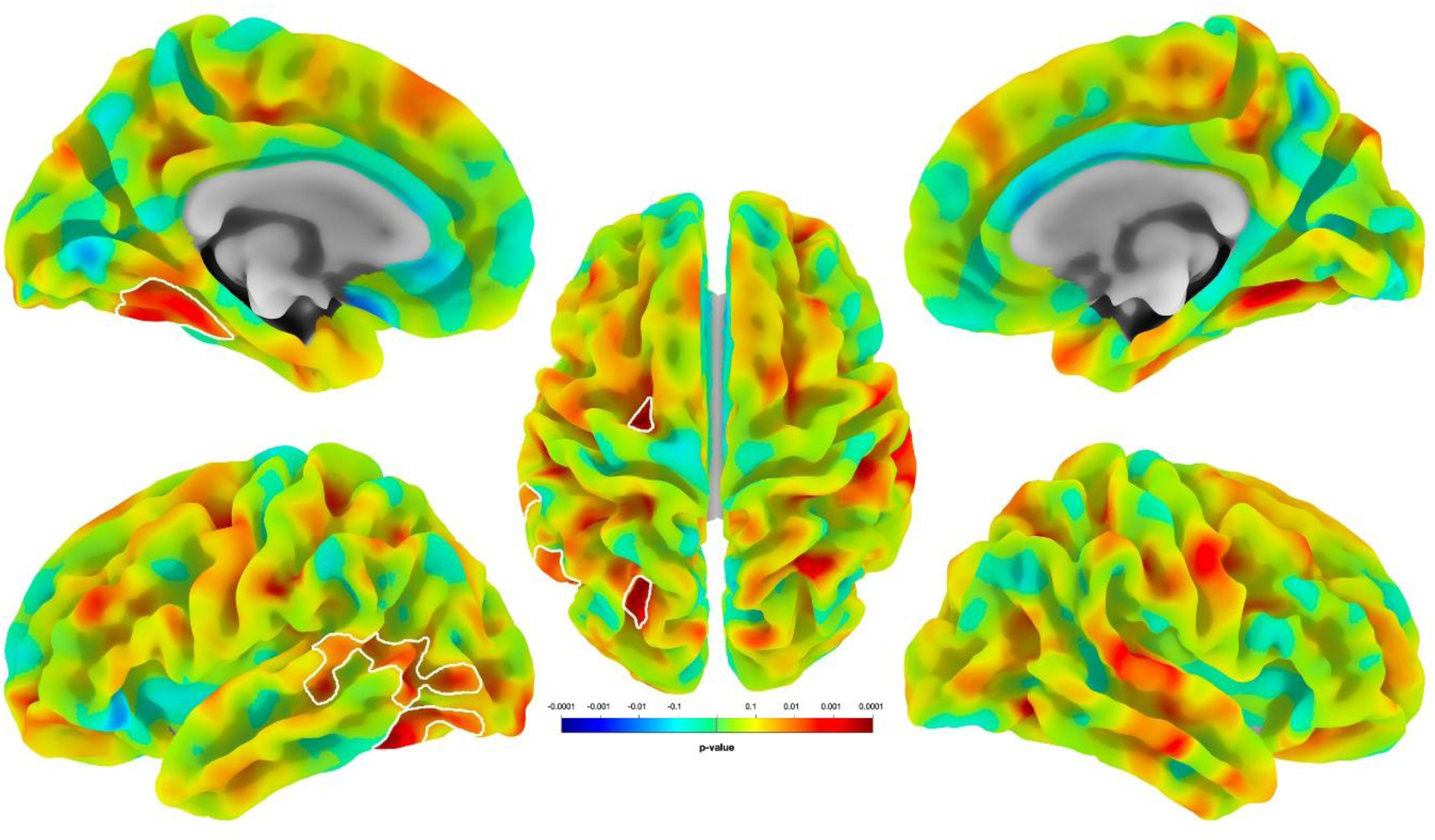
Regional Associations Between Cortical Thickness and Intraoperative Anesthetic-Dose Adjusted Fronto-Parietal EEG Alpha Band Power. Areas outlined in white represent areas with Threshold-Free Cluster Estimation p<0.05 for associations between cortical thickness and Intraoperative Anesthetic-Dose Adjusted FrontoParietal EEG Alpha Band Power. Hotter colors reflect areas in which cortical thinning was associated with lower Intraoperative Anesthetic-Dose Adjusted FrontoParietal EEG Alpha Band Power.

### ADA-FP alpha power is associated with POD timing and severity

POD data were available for 167 participants, and delirium severity could be calculated for 165. Median delirium severity was 2 [IQR: 1, 5], with a maximum observed score of 16. POD was detected in 77/167 participants (46.1%). Of these, 41 (53.2%) had POD detected only in the PACU, whereas 36 (46.8%) had POD detected at least once after PACU discharge, with or without concomitant PACU POD (Supplementary Table 3).

We next examined associations between intraoperative ADA-FP alpha power and POD incidence, timing, peak severity, and cumulative burden during hospitalization (Figure 5). There was a potential trend towards reduced intraoperative ADA-FP alpha power among participants who developed (vs those who did not develop) POD at any time point (either in the PACU and/or on the hospital ward/floor) that did not reach statistical significance (p=0.085; Figure 5A). ADA-FP alpha power did not differ significantly between participants without POD and those with PACU-only POD, whereas participants with ward POD had significantly lower ADA-FP alpha power than those without POD (P=0.019; Figure 5B). Lower intraoperative ADA-FP alpha power was associated with greater peak POD severity (Spearman’s ρ=-0.26, 95% bootstrap CI -0.401 to -0.107, P<0.001; Figure 5C) and with greater cumulative delirium burden during hospitalization, quantified as the area under the delirium severity-time curve (Spearman’s ρ=-0.28, 95% CI -0.42 to -0.13, P<0.001; Figure 5D).

**Figure 5.**
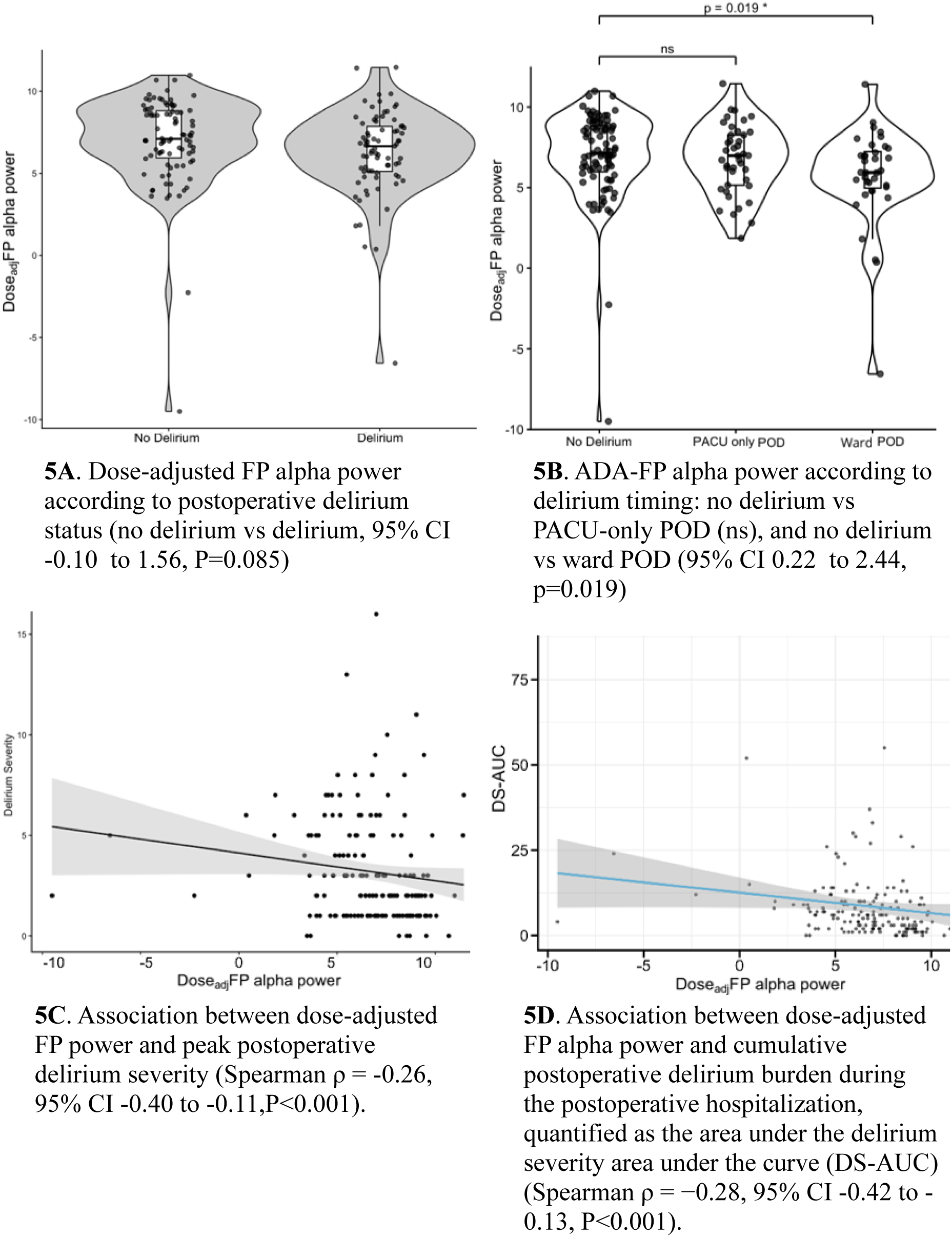
Relationships between intraoperative anesthetic dose-adjusted FP EEG alpha power and postoperative delirium incidence, peak severity and severity-time curve. Analyses included participants with available data for each delirium outcome. Violin plots show the distribution of dose-adjusted frontoparietal alpha power, with individual participant values overlaid. Each point represents one participant. ADA, Anesthesia Dose-adjusted; AUC, area under the curve; EEG electroencephalogram; FP frontoparietal; PACU, post-anesthesia care unit; POD, postoperative delirium.

## Discussion

Here, we demonstrate that lower ADA-FP alpha power is associated with altered preoperative functional connectivity within the DMN and cognitive control/DAN, cortical thinning within left temporoparietal and temporal regions, and increased rates of hospital ward POD, as well as greater POD severity and cumulative burden during the postoperative hospitalization (up to postoperative day 7). Numerous prior studies have linked lower intraoperative alpha power to preoperative cognitive impairment and POD, but the neuroanatomic basis of interindividual differences in anesthetic-induced alpha power remain unknown.(8–10,13,14) The findings presented here are novel and demonstrate that variation in ADA-FP alpha power is associated with preoperative differences in both cortical structure and intrinsic functional brain network connectivity.

These functional connectivity findings may provide insight into the underlying neurologic vulnerability that is reflected by low ADA-FP alpha power. Directing attention toward the external environment requires coordinated activity between large-scale networks, including the DMN, which supports internally directed cognition, and the DAN, which supports externally directed, goal-oriented attention.(50,51) We observed associations between ADA-FP alpha power and altered functional connectivity patterns involving portions of both the DMN and DAN. This raises the possibility that low ADA-FP alpha power reflects dysfunction within these functional brain networks that are key for switching from internally-focused to external attentional focus. Because inattention is a defining feature of delirium, this provides a plausible framework linking preoperative functional brain network connectivity dysfunction with an intraoperative EEG pattern (low alpha power) that is associated with both preoperative cognitive impairment and postoperative delirium. However, we did not directly test whether these connectivity patterns mediate the relationship between alpha power and POD, an important question to be addressed in future studies.

Additionally, the differences in functional connectivity within the DMN and DAN nodes seen here among patients with lower ADA-FP alpha power were essentially unchanged after controlling for the CSF Aβ42/40 ratio or CSF p-tau 217 levels, two different sensitive measures of Alzheimer’s Disease (AD) related amyloid pathology. This finding suggests that the DMN and DAN node functional connectivity alterations associated with lower ADA-FP alpha power in this cohort are not driven by pre-clinical/prodromal AD-related amyloid pathology. This raises an important question for future perioperative studies – what are the molecular and cellular mechanisms that are associated with (and/or which lead to) the alpha power-associated alterations in functional brain connectivity within the DMN and DAN observed here?

The cortical thickness findings provide complementary evidence that lower ADA-FP alpha power may reflect broader pre-existing brain vulnerability. Lower ADA-FP alpha power was associated with reduced cortical thickness across predominantly left-sided temporoparietal and temporal regions. Cortical thinning may reflect underlying neurodegenerative changes, including loss or shrinkage of neuropil and alterations in synaptic, neuronal, or glial architecture.(52) Prior work in older surgical patients has similarly linked reduced preoperative cortical thickness with greater postoperative delirium severity, supporting cortical thickness as a marker of pre-existing brain vulnerability. (53) Our findings extend this observation by demonstrating an association between cortical thinning specifically within left temporoparietal and temporal regions and lower ADA-FP alpha power. Whether cortical thinning within these brain regions causally or directly contributes to lower ADA-FP alpha power is unclear, particularly since frontoparietal alpha power is thought to reflect recurrent oscillations between the thalamus and frontoparietal cortex. Thus, it is unclear why cortical thinning within spatially distant regions (ie temporoparietal and temporal regions) would be associated with lower ADA-FP alpha power; it is also possible that cortical thinning in these brain regions is a marker or correlate but not a direct cause of lower ADA-FP alpha power.

We also found that ADA-FP alpha power was lower among participants who developed ward POD and that it was inversely associated with both peak delirium severity and cumulative delirium burden during the postoperative hospitalization. These findings are broadly consistent with prior studies linking lower intraoperative alpha power to POD and, more specifically, with our previous report in a different patient cohort, which showed that lower dose-adjusted frontoparietal alpha power was associated with both POD and moderate-to-severe delirium.(8,17) However, in the present cohort, ADA-FP alpha power was not significantly associated with overall binary POD incidence and did not differ between patients with PACU-only POD and those without POD. This differs from studies of emergence and recovery-room EEG, in which reduced alpha activity has been associated with PACU delirium.(54) These differences may reflect the distinct EEG phenotypes being measured: our metric summarizes ADA-FP alpha power during the maintenance phase of general anesthesia, whereas prior PACU studies examined EEG during emergence from anesthesia or postoperative recovery. Together, these findings suggest that the relationship between perioperative alpha activity and POD may depend not only on whether delirium occurs, but also on when alpha activity is measured-ie during the maintenance vs emergence phases of general anesthesia. The fact that lower ADA-FP alpha power was associated with postoperative hospital ward delirium but not with PACU delirium may mean that there are differing neurobiological correlates and perhaps even different mechanisms that underlie delirium that presents during these two different phases of postoperative care. Alternatively, the lack of difference in ADA-FP alpha power between patients with PACU delirium vs no delirium may reflect an insufficient sample size in this study (ie a lack of statistical power).

Prior work has supported the concept that low alpha power may represent a vulnerable neurophysiologic response to anesthesia.(13) Existing mediation studies suggest that altered intraoperative EEG response may partially mediate associations between preoperative vulnerability and POD.(55) Our findings extend this concept by showing that lower ADA-FP alpha power is associated with preoperative structural and functional brain differences as well as with ward POD, and greater overall POD delirium severity and the cumulative postoperative POD severity burden. These observations are consistent with the broader view that POD reflects an interaction between pre-existing brain vulnerability and perioperative stressors, rather than anesthetic exposure alone.(4)

### Several limitations should be considered

First, this was a single-center study conducted at a U.S. academic medical center; the generalizability of the findings to other surgical populations, anesthetic practices, and clinical settings is unclear. Thus, it will be important to conduct studies to replicate these findings at other centers and in other patient populations. Second, the multimodal data-quality requirements excluded participants with incomplete or lower quality EEG or MRI data, resulting in a smaller sample than originally enrolled. Nonetheless, the cohort described here was still of sufficient size to identify links between structural and functional neuroimaging, intraoperative anesthetic brain responses, and postoperative delirium. Third, this cohort also included relatively few participants at the lowest end of the intraoperative alpha-power distribution;(10) nonetheless, this cohort had sufficient variance in intraoperative ADA FP alpha power to identify associations with neuroimaging and delirium phenotypes. Fourth, resting-state fMRI measures correlations in the blood oxygen level dependent signal rather than neuronal activity directly, and the observed relationships between preoperative connectivity and intraoperative alpha power should not be interpreted as direct measures of correlated neural activity. Future mechanistic studies using methods that more directly capture neuronal activity, such as intracranial electrocorticography (ECoG) or magnetoencephalography (MEG), could help determine whether the functional MRI connectivity correlates of low ADA-FP alpha power identified here reflect actual correlated electrical activity changes in these brain regions. Fifth, although delirium was assessed repeatedly in the PACU and during hospitalization, delirium is an intrinsically fluctuating syndrome, and transient episodes between scheduled assessments may have been missed. Nonetheless, the postoperative ward delirium rates seen here match those seen in other large delirium studies,(48) and a sufficient number of delirium cases were seen to identify associations with intraoperative EEG alpha power. Sixth, because artifactual EEG channels contributed zero EEG power to ROI averages when other channels in the ROI were usable, ROI-specific alpha power may have been modestly underestimated during periods of partial artifact. Variable mastoid electrode availability may have introduced additional measurement variability. Despite this potential issue, the present data were nonetheless sufficient to find associations between EEG alpha power and both neuroimaging features and postoperative delirium.

### This study also has several significant strengths

This prospective study incorporated multimodal phenotyping that included preoperative structural MRI and resting-state functional MRI, CSF biomarker measurements, intraoperative 32-channel EEG recordings, perioperative real-life anesthetic and surgical exposure, and repeated POD assessments using state of the art assessments. The use of 32-channel intraoperative EEG enabled regional rather than single-electrode estimates of frontoparietal alpha activity. Averaging across anatomically defined electrode groups also reduces the influence of noise or poor signal quality from individual channels and provides a more stable representation of regional alpha activity, while retaining broader frontoparietal information across the scalp. Rigorous EEG preprocessing included artifact detection, exclusion of periods affected by potentially confounding anesthetic and adjunct medications, and minute-level adjustment for the volatile anesthetic exposure. Stringent rs-fMRI quality-control procedures were also applied. To avoid the influence of motion artifact within the resting state data, subject-level motion correction was set conservatively at >0.5mm (i.e., excludes volumes with framewise displacement of >0.5mm) with additional group-level correction for motion in all analyses.(56) Moreover, resting-state functional imaging was acquired using the Inscapes paradigm, a low-demand abstract audiovisual stimulus designed to improve participant compliance while minimizing cognitive engagement.(24) Finally, repeated delirium assessments in the PACU and throughout hospitalization allowed us to assess postoperative delirium incidence, peak severity, and the cumulative burden of in-hospital delirium following surgery.

Overall, the findings presented here both 1) replicate and extend the existing literature linking reductions in intraoperative anesthetic induced EEG alpha power to increased delirium risk and severity, and 2) identify functional and structural neurologic alterations within key regions of the brain’s default mode network and dorsal attention networks that underlie reductions in intraoperative anesthetic induced EEG alpha power among older surgical patients. More broadly, the combination of structural and functional neuroimaging, CSF biomarker assays, intraoperative EEG recordings and postoperative delirium assessments reported here provide a multimodal framework for dissecting the underlying mechanisms of perioperative neurocognitive disorders.

## Data Availability

All data produced in the present study are available upon reasonable request to the authors

## Acknowledgments

This work was supported by the National Institutes of Health, National Institute on Aging (grant R01-AG073598), and by the Stanford University Department of Anesthesiology, Perioperative and Pain Medicine. The authors acknowledge the Stanford Human Immune Monitoring Center (HIMC) for performing the NULISA assays.

