## Supplemental Tables 1-3 Figures 1-2 for "Functional and Structural Neuroanatomic Correlates of Intraoperative Anesthetic Dose-Adjusted EEG Alpha Power Among Older Surgical Patients"

Supplementary Table 1: Intraoperative Anesthetic & Surgical Characteristics

| Perioperative factors | n=168 |
|---|---|
| Surgical specialty | |
| Thoracic | 34 (20.2%) |
| General Surgery | 55 (32.7%) |
| Gynecology | 12 (7.1%) |
| Neurosurgery non-cranial | 11 (6.5%) |
| Obstetrics | 1 (0.6%) |
| Orthopedics | 13 (7.7%) |
| Plastic Surgery | 3 (1.8%) |
| Urology | 39 (23.2%) |
| Surgical Duration (min) | 214 (132) |
| Volatile anesthetic agent | |
| Isoflurane | 62 (36.9%) |
| Sevoflurane | 106 (63.1%) |
| aaMAC | 0.9 (0.2) |
| Estimated Blood Loss (mL) | 97 (313) |

Data are presented as n (%), mean (SD), or median [25th, 75th percentiles], as appropriate aaMAC, age-adjusted minimum alveolar concentration.

Supplementary Table 2: Seed-to-Voxel Analysis of Brain regions that show differential functional connectivity relationships with MVPA-enumerated seed regions from Figure 2, in association with intraoperative anesthetic dose adjusted fronto-parietal alpha power.

| | Anatomical Location | Cluster maxima (x, y, z) | Cluster extent (k) | Cluster p-FWE | Functional Association |
|---|---|---|---|---|---|
| Cluster 1 (B.1) | R Lateral Occipital Cortex | (40, -62, -10) | 2021 | <0.001 | Visual |
| | L Lateral Occipital Cortex | (-28, -84, -4) | 1510 | <0.001 | Visual |
| | R Frontal Pole | (36, 48, 0) | 767 | <0.001 | Cognitive Control |
| | R Angular/ Supramarginal Gyrus | (60, -48, 44) | 634 | <0.001 | Cognitive Control |
| | Anterior Cingulate | (10, 26, 44) | 495 | <0.001 | Cognitive Control |
| | L Angular/ Supramarginal Gyrus | (-62, -56, 32) | 234 | 0.022 | Default Mode |
| | Posterior Cingulate | (2, -28, 46) | 218 | 0.030 | Default Mode |
| Cluster 2 (B.2) | L Post/Pre-central Gyrus | (-56, -10, 32) | 1278 | <0.001 | Somatosensory Motor |
| | R Post/Pre-central Gyrus | (62, -2, 26) | 1273 | <0.001 | Somatosensory Motor |
| | Posterior Cingulate | (2, -34, 20) | 852 | <0.001 | Default Mode |
| | R Frontal Pole | (20, 64, 4) | 489 | <0.001 | Cognitive Control |
| | R Superior Temporal Gyrus | (60, 4, -6) | 459 | <0.001 | Somatosensory Motor |
| | L Temporal Pole | (-52, 8, -8) | 320 | 0.006 | Default Mode |
| | R Frontal Pole | (14, 32, 62) | 309 | 0.007 | Cognitive Control |
| | R Cerebellum* | (46, -66, -54) | 216 | 0.034 | - |
| | R Supplementary Motor Cortex | (2, 10, 50) | 214 | 0.035 | Salience/Ventral Attention |
| | R Middle/Inferior Temporal Gyrus | (68, -34, -24) | 195 | 0.050 | Cognitive Control |
| Cluster 3 (B.3) | R Lateral Occipital Cortex | (44, -84, -14) | 800 | <0.001 | Visual |
| | R Lateral Occipital Cortex | (36, -72, 2) | 602 | <0.001 | Visual |
| | R Frontal Pole | (24, 48, 6) | 538 | <0.001 | Cognitive Control |
| | L Frontal Pole | (-40, 48, 8) | 383 | 0.002 | Cognitive Control |
| | Anterior Cingulate | (4, 36, 20) | 330 | 0.004 | Cognitive Control |
| | L Superior Frontal Gyrus | (-24, 8, 68) | 318 | 0.005 | Dorsal Attention |
| Cluster 4 (B.4) | R Angular/ Supra-marginal Gyrus | (46, -46, 32) | 570 | <0.001 | Default Mode |
| | L Angular/ Supra-marginal Gyrus | (-54, -54, 32) | 563 | <0.001 | Default Mode |
| | R Middle Temporal Gyrus | (62, -22, -12) | 435 | <0.001 | Default Mode |

* The Schaefer atlas(49) does not provide a functional network assignment for this region.

Supplementary Table 3: Postoperative delirium incidence and timing in the EEG analytic cohort

| Characteristic | n/N (%) |
|---|---|
| Participants with postoperative delirium data | 167 (100%) |
| No postoperative delirium | 90/167 (53.9%) |
| Postoperative delirium | 77/167 (46.1%) |
| - POD diagnosed in the PACU only | 41/77 (53.2%) |
| - POD diagnosed in the ward, with or without PACU POD | 36/77 (46.8%) |

EEG, Electroencephalogram; PACU, post-anesthesia care unit; POD, postoperative delirium.

Supplementary Figure 1. Preoperative resting-state functional connectivity associations with intraoperative dose-adjusted frontoparietal alpha power after adjustment for CSF Aβ42/Aβ40 ratio, shown across eight views

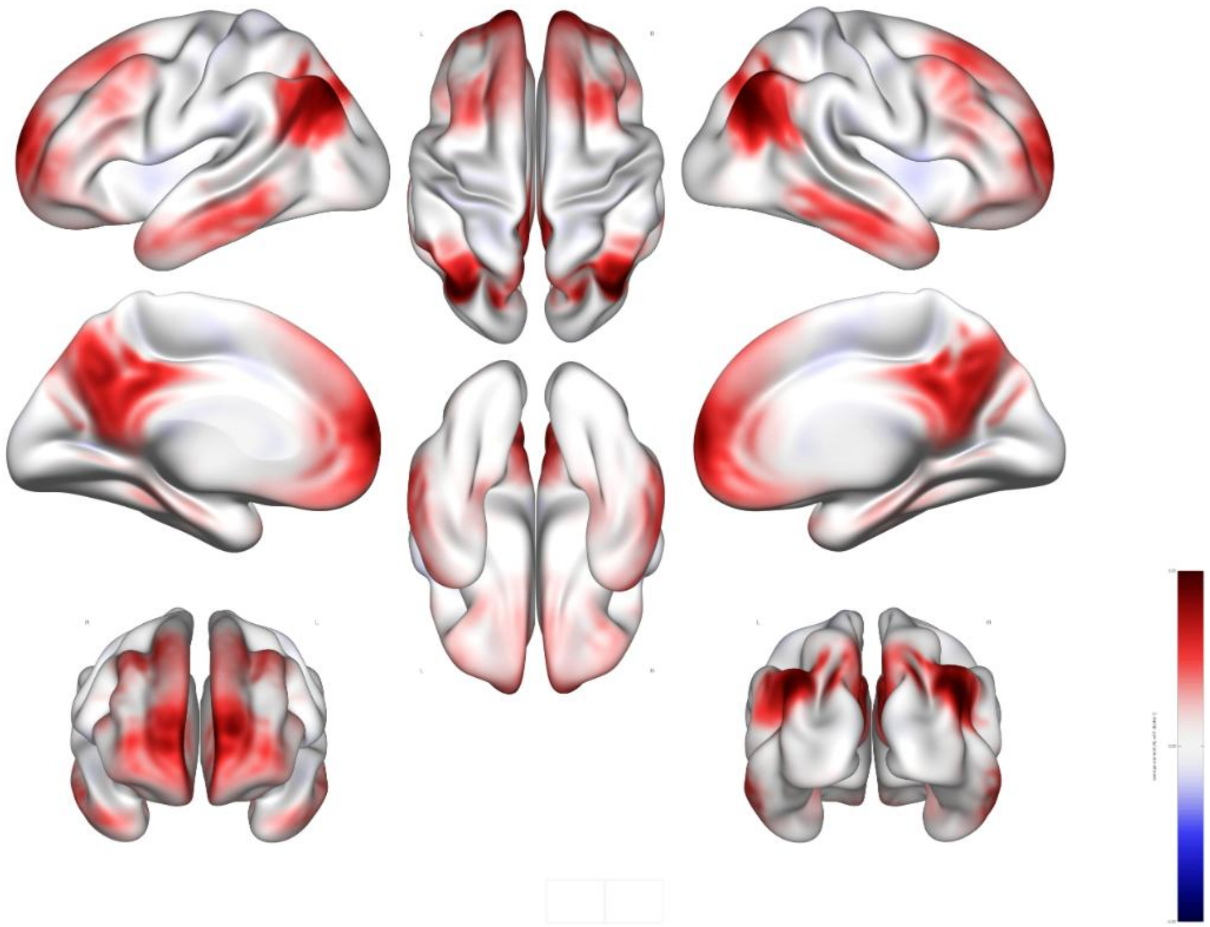

Supplementary Figure 2. Preoperative resting-state functional connectivity associations with intraoperative dose-adjusted frontoparietal alpha power after adjustment for CSF p-tau217 levels, shown across eight views

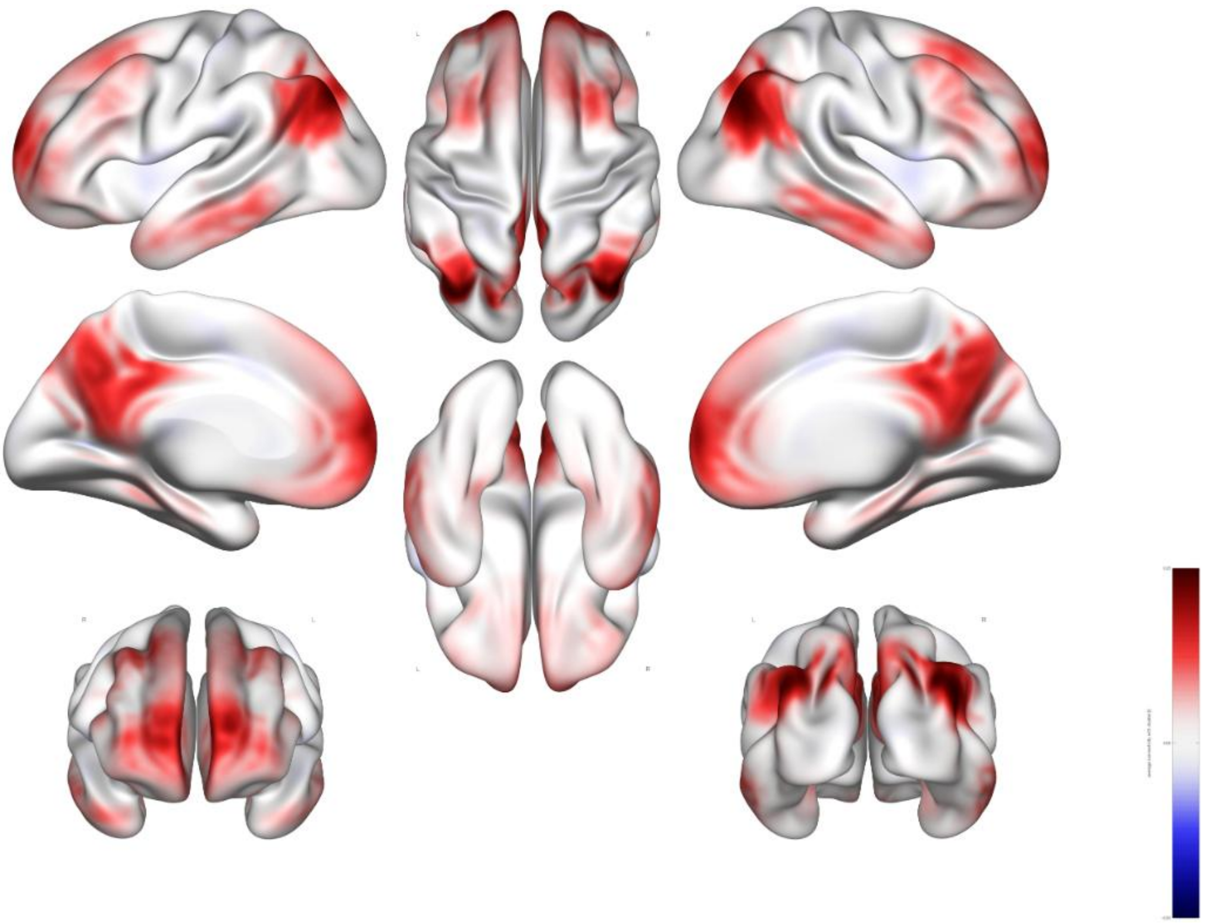
